# Exome sequencing identifies *HELB* as a novel susceptibility gene for non-mucinous, non-high-grade-serous epithelial ovarian cancer

**DOI:** 10.1101/2024.04.02.24304968

**Authors:** Ed M. Dicks, Jonthan P. Tyrer, Suzana Ezquina, Michelle Jones, John Baierl, Pei-Chen Peng, Michael Diaz, Ellen Goode, Stacey J Winham, Thilo Dörk, Toon Van Gorp, Ana De Fazio, David Bowtell, Kunle Odunsi, Kirsten Moysich, Marina Pavanello, Ian Campbell, James D. Brenton, Susan J. Ramus, Simon A. Gayther, Paul D. P. Pharoah

## Abstract

Rare, germline loss-of-function variants in a handful of genes that encode DNA repair proteins have been shown to be associated with epithelial ovarian cancer with a stronger association for the high-grade serous hiostotype. The aim of this study was to collate exome sequencing data from multiple epithelial ovarian cancer case cohorts and controls in order to systematically evaluate the role of coding, loss-of-function variants across the genome in epithelial ovarian cancer risk. We assembled exome data for a total of 2,573 non-mucinous cases (1,876 high-grade serous and 697 non-high grade serous) and 13,925 controls. Harmonised variant calling and quality control filtering was applied across the different data sets. We carried out a gene-by-gene simple burden test for association of rare loss-of-function variants (minor allele frequency < 0.1%) with all non-mucinous ovarian cancer, high grade serous ovarian cancer and non-high grade serous ovarian cancer using logistic regression adjusted for the top four principal components to account for cryptic population structure and genetic ancestry. Seven of the top 10 associated genes were associations of the known ovarian cancer susceptibility genes *BRCA1, BRCA2, BRIP1, RAD51C, RAD51D, MSH6* and *PALB2* (false discovery probability < 0.1). A further four genes (*HELB, OR2T35, NBN* and *MYO1A*) had a false discovery rate of less than 0.1. Of these, *HELB* was most strongly associated with the non-high grade serous histotype (P = 1.3x10^−6^, FDR = 9.1x10^−4^). Further support for this association comes from the observation that loss of function variants in this gene are also associated with age at natural menopause and Mendelian randomisation analysis shows an association between genetically predicted age at natural menopause and endometrioid ovarian cancer, but not high-grade serous ovarian cancer.

## Introduction

Substantial progress has been made in the past 30 years in identifying inherited genetic variation associated with an increased risk of epithelial ovarian cancer. The “high-penetrance” genes *BRCA1* and *BRCA2* were identified by linkage studies in the 1990’s; protein truncating variants in these genes confer a substantial lifetime risk of epithelial ovarian cancer as well as breast cancer and other cancers. Epithelial ovarian cancer is also known to be part of the Lynch Syndrome phenotype associated with protein truncating variants in the mis-match repair genes. Rare coding variants in *BRIP1, PALB2, RAD51C* and *RAD51D* have been shown to confer more moderate risks by using candidate-gene case-control sequencing ^1-3^. Also, over the past 15 years, large-scale genome-wide association studies (GWAS) conducted by the Ovarian Cancer Association Consortium (OCAC) have identified more than 40 common susceptibility alleles ^4^.

There are five major histotypes of epithelial ovarian cancer - high-grade serous, low-grade serous, clear cell, endometriod and mucinous - which share a substantial fraction of their heritable component ^5^. Nevertheless, there are some notable differences in their genetic risk factors. High- and moderate-penetrance risk variants in *BRCA1, BRCA2, BRIP1, RAD51C* and *RAD51D* predispose to high-grade serous EOC whereas loss-of-function variants in the mis-match repair genes predispose to endometrioid and clear cell EOC. There are also histotype specific differences in the risks conferred by common risk alleles with the mucinous histotype in particular being substantially different from the other histotypes ^4^.

The uncommon and rare, high- and moderate penetrance alleles identified to date explain about one quarter of the inherited component of epithelial ovarian cancer susceptibility with a further 5 percent explained by the known common risk alleles. Genome-wide heritability analyses have estimated that the set of common variants that are tagged or captured by the standard genome-wide genotyping arrays explains about 40 percent of the familial aggregation –the so-called chip heritability. The characteristics of the alleles that account for the remaining familial aggregation are not known; analyses of whole-genome data suggest that a substantial portion is explained by rare variants. Only a small fraction of genes, mostly those involved in DNA repair, have been examined for risk association using the large sample sizes needed to detect modest risks. Hence, there could be many more genes conferring similar risks yet to be discovered. The aim of this project was to identify genes with rare coding variants that confer loss of function (LoF) that are associated with risk of epithelial ovarian cancer.

## Methods

### Description of case and control datasets

Germline whole exome sequencing (WES) data and whole genome sequencing (WGS) data as BAM or CRAM files from multiple epithelial ovarian cancer case series were collated from multiple sources (Table 1). Control sequencing data were sourced wholly from the UK Interval study; a set of healthy UK blood donors (https://www.intervalstudy.org.uk/). All analyses restricted case histotypes to high grade serous, low grade serous, clear-cell, endometrioid, mixed, and other rare histotypes. Mucinous ovarian cancer cases were excluded because it has previously been shown that the genetic etiology of this histotype differs substantially from the other histotypes. In total, exome or whole genome sequencing data were available for 1,638 cases and 4,502 controls. We also used the variant calls (as VCF files) for 1,099 EOC cases and 9,423 cancer free controls from the WES sequencing released by UK Biobank (UKB) (https://www.ukbiobank.ac.uk/). Cases were individuals with a diagnosis of invasive epithelial ovarian cancer (ICD10 code C56) with clear cell, endometrioid, papillary, other and serous histology codes. Controls were age matched women without a cancer diagnosis and without a history of oophorectomy. Up to ten controls were selected for each case. Thus, the final sample size was 2,573 cases and 13,925 controls before quality control. All samples were from studies approved by a local ethical committee and all participants gave written informed consent.

**Table 1:** Number of non-Biobank epithelial ovarian cancer patients by source of sequencing (total before sample QC 1,638; total after QC 1,474)

| Data source | Case type | Number<br>(before/after QC) | Reference | Accession number |
| --- | --- | --- | --- | --- |
| *Exomes* |  |  |  |  |
| UK Familial Ovarian Cancer Registry | BRCA1/2-negative with family history | 53/42 | Unpublished |  |
| Leuven | Unselected | 45/44 | Unpublished |  |
| Gilda-Radner Ovarian Cancer Registry | BRCA1/2-negative with family history | 96/80 | Unpublished |  |
| OCAC | Positive family history or <50 years of age | 262/232 | Unpublished |  |
| Mayo Clinic | Unselected | 25/24 | Unpublished |  |
| Campbell | BRCA1/2-negative high-grade serous | 536/493 | <sup>6</sup> | EGAD00001006030 |
| Hannover | Unselected | 11/7 | Unpublished |  |
| TCGA | High grade serous | 413/361 |  | phs000441.v2.p6 |
| *Whole genomes* |  |  |  |  |
| Peter MacCallum Cancer Centre | High-grade serous | 93/92 | <sup>7</sup> | EGAC00001000010 |
| Bowtell | Long term survivors with high-grade serous | 48/46 | <sup>8</sup> | EGAC00001002510 |
| BRITROC | High grade serous | 56/53 | <sup>9</sup> | EGAD00001004189 |

### Variant calling and filtering

All BAM/CRAM files were aligned to human genome version hg19/GRCh37. The original TCGA EOC BAM files had been aligned against human genome hg18/NCBI36, these data were lifted over to build hg19 with the CrossMap s/w ^10^ to match the rest of the WES/WGS data. All non-Biobank sequencing data were analysed in an identical way. Duplicate sequence reads were removed with the *picard* sequencing tools ^11^. Sequence reads were partitioned per chromosome and general manipulation performed with the samtools s/w ^12^. Variants were called with the Genome Analysis ToolKit (GATK) UnifiedGenotyper version 3.8-1 ^13^, and following the best practices guide as appropriate to our data ^14,15^. We restricted our risk variant discovery to substitutions and short indels (length <= 12 bp). Variants were annotated with *ANNOVAR* ^16^ referred to the UCSC RefSeq gene transcript set (https://genome.ucsc.edu). Protein coding transcripts with an NM_^*^ type identifier were used, with the transcript having the longest coding sequence being chosen for genes with multiple transcripts. This yielded 19,092 gene transcripts at human reference hg19/GRCh37 for variant annotation. The averaged coverage of targeted bases at 10X for the non-UKB samples was 91 percent for cases and 92 percent for controls.

UK Biobank VCF calls were based on build GRCh38 ^17^; in order to incorporate these data into our pipeline we lifted over these calls to build GRCh37 using *picard* and inserted the data at the appropriate step. Only Biobank VCF calls with depth (DP) greater than or equal to 10 and genotyping quality (GQ) greater than or equal to 20 were retained.

Variant calls from GATK were filtered with an in-house hard filter tuned for optimum sensitivity by comparison of WES calls with chip genotyping calls from multiple genotyping arrays [see Chip Genotyping Data for details]. Additionally for all call sets, variants were carried forward only if depth (DP) >= 10 and alternate allele frequency (AAF) >= 15 percent. A more stringent filter was also applied, assigning calls a high quality (HQ) having AAF >= 20 percent and number of alternate alleles >= 4. All variant sites with at least 1 occurrence of an HQ call were retained, whilst sites without any HQ calls were rejected.

Rare variants, those with minor allele frequency (MAF) <= 1 percent, were visually inspected with the Integrative Genomics Viewer (IGV) software ^18^ and rejected if any doubts raised, e.g. not called bidirectionally. Visual inspection of variants called for UK Biobank was only carried out for those variant sites not validated in the non-Biobank data.

QC was applied to each rare variant site, rejecting sites with genotype frequencies showing significant deviation from those expected under Hardy-Weinberg equilibrium in either cases or controls (p-value < 1e-15 and for UKB and non-UKB separately), and those with missingness > 20 percent (proportion of samples with depth<10). We also tested each variant for association with epithelial ovarian cancer and excluded those with minor allele frequency <0.01 and test for association for variant p-value <= 1e-6; association p-value <= 1e-7 and 0 rare control alleles; and association p-value <= 1e-10. Pre-QC filtering was also applied to the TCGA calls to check for inconsistency with non-TCGA, since TCGA contained a higher rate of false positive calls.

### Variant classification

Variants were defined as loss-of-function according to the following criteria: 1) Variants predicted to cause protein truncation, that is stopgain variants, frameshift indels, and canonical splice site variants. 2) Non-canonical splice site variants and in-exon variation within 3 bp of the exon-splice boundary predicted by the MaxEntScan algorithm to disrupt splicing ^19^. Qualifying variants with a wild-type MaxEntScan score greater than or equal to 3 and decreased by greater than 40 percent in comparison to the reference sequence were assumed to be deleterious. 3) Missense single nucleotide variants or in frame indels designated by multiple submitters to the NCBI Clinvar database (https://www.ncbi.nlm.nih.gov/clinvar) as either pathogenic or likely pathogenic with no conflicts between submitters.

### Sample quality control and exclusion

Samples were removed if they met any of the following criteria: i) low average depth of coverage (< 25 percent at 10X) ii) excess LoF calls (>1000) iii) concordance of exome data variant calls and chip genotyping calls (see below) of < 97 percent iv) known duplicates or cryptic duplicate sample based on common variant calls. After exclusions, a total of 1,474 cases and 4,500 controls remained in the non-Biobank set and 1,099 cases and 9,423 controls in the Biobank set (Table 1 and Table 2).

**Table 2:** Number of epithelial ovarian cancer patients by histotype after QC.

| Histotype | Non-UKB | UKB | Total | % of total |
| --- | --- | --- | --- | --- |
| High grade serous* | 1,209 | 667 | 1876 | 72.9 |
| Low grade serous | 51 | 0 | 51 | 2.0 |
| Clear cell | 52 | 86 | 138 | 5.4 |
| Endometrioid | 110 | 139 | 249 | 9.7 |
| Mixed | 19 | 5 | 24 | 0.9 |
| Undifferentiated | 16 | 27 | 43 | 1.7 |
| Other | 17 | 175 | 192 | 7.5 |

\* includes those with carcinosarcoma and serous carcinoma of unspecified grade

### Chip genotyping data

We used chip genotype calls to tune filters for raw variant calling and check integrity of sample naming and also as an additional level of QC for any non-UKB samples overlapping chip manifests. Data were from four different Ovarian Cancer Association Consortium genotyping projects (OncoArray ^4^, iCOGS ^20^, exome chip ^21^, and an ovarian GWAS ^22^), and TCGA. The numbers of WES/WGS samples overlapping with each chip were 323, 350, 81, 95, and 412, respectively.

### Association analysis

We carried out a gene-by-gene simple burden test for association of rare loss-of-function variants with all non-mucinous ovarian cancer and high grade serous ovarian cancer. Rare variants were defined as those with a minor allele frequency of less than 0.1 percent. We classified each individual for each gene as a loss-of-function variant carrier or non-carrier, depending on whether they had at least one rare variant (below the MAF threshold) in that gene or not. Then we performed a logistic regression for each gene adjusting for the top four principal components to account for cryptic population structure and genetic ancestry. Principal component analysis for the non-UK Biobank data was carried out using data from 36,047 uncorrelated variants (pairwise r2 < 0.1) with MAF >0.03 using an in-house program (available at http://ccge.medschl.cam.ac.uk/software/pccalc/). Principal components for the UK Biobank samples were provided by UK Biobank ^23^. We also adjusted for study stratum - non-UKB, UKB 50K sample set and UKB non-50K sample set. The UK Biobank data were stratified on recommendation from UK Biobank, since different oligo lots had been used in the 2 stages of UK Biobank sample sequencing.

We calculated a false discovery probability based on the methods of Benjamini and Hochberg ^24^ and a Bayes False Discovery Probability using the method proposed by Wakefield ^25^. For the latter method we assumed a prior probability of association for any one gene of 0.005 – ∼100 expected true number of genes truly associated with epithelial ovarian cancer – and a likely maximum effect size (log odds ratio) of 0.836.

## Results

An initial analysis was performed for all genes in the non-UKB set of 1,474 cases and 4,500 controls that passed QC. There were 12,761 genes with at least one case or control loss-of-function variant carrier with minor allele frequency of less than 0.1 percent, of which 4623 had sufficient pathogenic variant carriers to obtain a risk estimate. There was little evidence of inflation of the test statistic (Figure 1). Seven hundred and thirty-seven genes had a P-value for association of less than 0.05; these genes were selected for additional analysis in the UKB data in addition to candidate genes *ATM, BARD1, CHEK2, FANCM, MLH1, MSH2, MSH6, PMS2, RAD51B, SLX4, TIPARP* and *TP53*.

**Figure 1:**
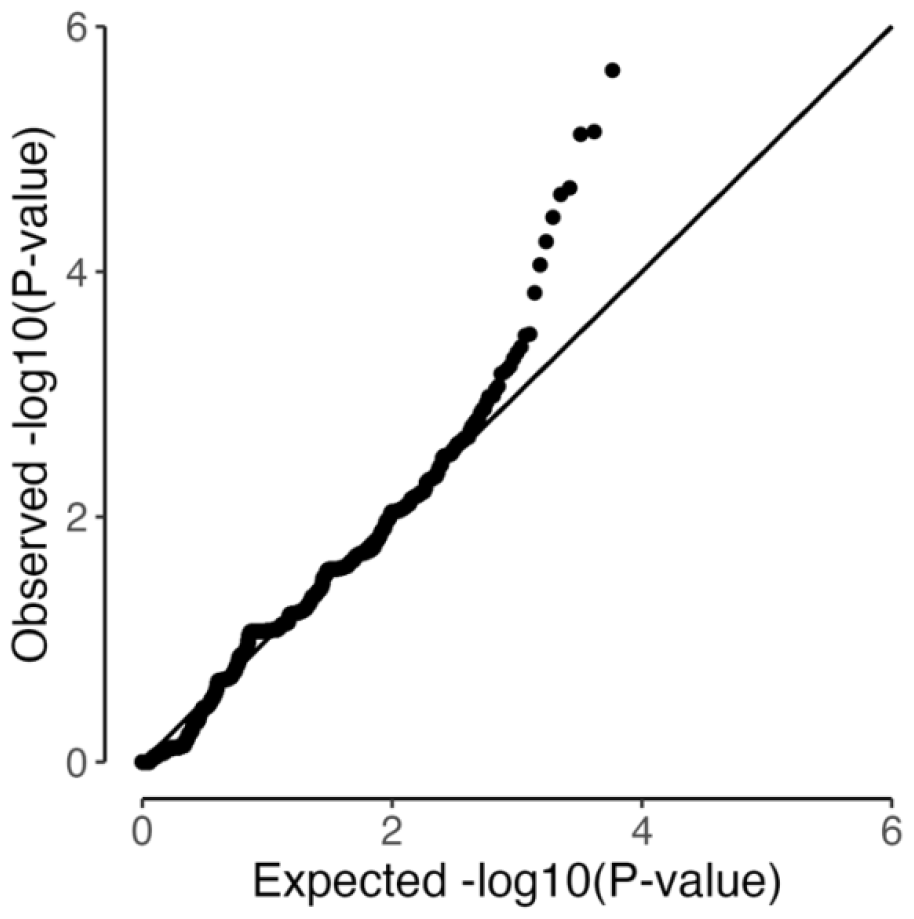
QQ plot for association analysis of genes with at least one case or control loss-of-function variant carrier in the non-UK Biobank data set (three genes with smalles P-values excluded)

The results of the association analysis for each gene with non-mucinous ovarian cancer, high-grade serous ovarian cancer and non-high grade serous ovarian cancer are shown in Supplementary Table 1. The association with the smallest P-value from the three histotype specific analyses was selected for each gene and the genes were then ranked by P-value. Table 3 shows the top 20 associated genes based on ranked Bayes False Discovery Probability. Seven of the top 10 associated genes were the known ovarian cancer susceptibility genes *BRCA1, BRCA2, BRIP1, RAD51C, RAD51D, MSH6* and *PALB2* (all with a False Discovery Rate < 0.1). A further four genes (*HELB, OR2T35, NBN* and *MYO1A*) had a False Discovery Rate of less than 0.1 with three more genes (*NENF, MIGA1, STARD6*) having a False Discovery Rate of less than 0.2. *BRCA1, BRCA2, BRIP1, RAD51C, RAD51D, PALB2, NENF, MIGA1 and STARD6* were more strongly associated with high-grade serous ovarian cancer whereas *HELB, OR2T35, NBN, MSH6* and *MYO1A* were more strongly associated with the non-high grade serous histotypes (Supplementary Table 2).

**Table 3:** Genes most strongly associated with epithelial ovarian cancer.

| Case type | Gene | Minor allele freq | Odds ratio | (95% CI) | P-value | FDR | BFDP |
| --- | --- | --- | --- | --- | --- | --- | --- |
| HGSOC | *BRCA2* | 0.0036 | 12 | (8.3 – 18) | 6.6x10<sup>-38</sup> | 6.8x10<sup>-34</sup> | 3.6x10<sup>-32</sup> |
| HGSOC | *BRCA1* | 0.0010 | 39 | (22 - 70) | 7.0x10<sup>-36</sup> | 3.6x10<sup>-32</sup> | 1.1x10<sup>-28</sup> |
| HGSOC | *BRIP1* | 0.0011 | 11 | (5.4 - 20) | 3.4x10<sup>-12</sup> | 5.8x10<sup>-9</sup> | 3.2x10<sup>-7</sup> |
| HGSOC | *RAD51C* | 0.00057 | 12 | (4.9 - 29) | 5.2x10<sup>-8</sup> | 6.7x10<sup>-5</sup> | 0.0031 |
| NHGSOC | *HELB* | 0.00093 | 11 | (4.3 – 31) | 3.3x10<sup>-6</sup> | 0.0014 | 0.052 |
| HGSOC | *RAD51D* | 0.00043 | 13 | (4.3 - 36.4) | 3.3x10<sup>-6</sup> | 0.0030 | 0.12 |
| NHGSOC | *NBN* | 0.0015 | 8.6 | (3.2 - 23.1) | 2.2x10<sup>-5</sup> | 0.016 | 0.29 |
| NHGSOC | *OR2T35* | 0.00072 | 15 | (4.4 - 48.7) | 1.1x10<sup>-5</sup> | 0.009 | 0.31 |
| NHGSOC | *MSH6* | 0.0013 | 7.9 | (2.9 - 21.2) | 4.5x10<sup>-5</sup> | 0.028 | 0.41 |
| HGSOC | *PALB2* | 0.0017 | 3.9 | (2 - 7.8) | 9.7x10<sup>-5</sup> | 0.058 | 0.42 |
| NHGSOC | *MYO1A* | 0.0024 | 10 | (3.2 - 28.2) | 4.3x10<sup>-5</sup> | 0.028 | 0.45 |
| HGSOC | *NENF* | 0.00050 | 7.8 | (2.7 - 23.1) | 1.8x10<sup>-4</sup> | 0.10 | 0.69 |
| HGSOC | *STARD6* | 0.00050 | 7.1 | (2.4 - 20.8) | 3.7x10<sup>-4</sup> | 0.18 | 0.79 |
| HGSOC | *MIGA1* | 0.00022 | 15 | (3.5 - 65.2) | 2.6x10<sup>-4</sup> | 0.14 | 0.85 |
| HGSOC | *OR4A47* | 0.00036 | 8.5 | (2.5 - 29.3) | 6.6x10<sup>-4</sup> | 0.26 | 0.87 |
| NMOC | *PLEKHG5* | 0.0019 | 3.9 | (1.7 - 9) | 0.0014 | 0.42 | 0.88 |
| HGSOC | *LIPT1* | 0.00050 | 5.4 | (2 - 15) | 0.0011 | 0.38 | 0.88 |
| HGSOC | *ACADM* | 0.0015 | 3.4 | (1.6 - 7.3) | 0.0020 | 0.49 | 0.90 |
| NMOC | *DQX1* | 0.0017 | 3.2 | (1.5 - 6.7) | 0.0022 | 0.49 | 0.90 |
| NMOC | *IFT172* | 0.0012 | 3.5 | (1.6 - 7.9) | 0.0023 | 0.49 | 0.91 |

FDR – Benajmini-Hochberg false discovery rate
BFDP – Bayes false discovery probability
HGSOC – high grade serous ovarian cancer
NMOC – non-mucinous epithelial ovarian cancer
NHGSOC –non-high grade serous ovarian cancer (non-mucinous)

## Discussion

We have assembled whole exome sequencing to generate a large epithelial ovarian cancer case-control study in order to investigate the role of rare, loss-of-function coding variation in the germline and risk of epithelial ovarian cancer. The exome sequencing of the non-UK Biobank cases and controls was carried out in different centres with the potential for false positive associations that are due to technical artefacts resulting in differential variant calls between cases and controls. We attempted to limit such bias by harmonising the variant calling across the different data sets with careful visual inspection of many variants using the Integrative Genomics Viewer. The lack of inflation of the test statistics for the gene-based association tests within the non-UK Biobank data suggests that any technical bias was small (if present).

Perhaps the major limitation of this study was the limited power to detect rare variants with modest effects. Figure 2A shows the power of the available sample size to detect loss-of-function alleles by carrier frequency and effect size. Power to detect alleles with effects similar to the known genes is good, but power to detect alleles conferring odds ratios between 2 and 5 is limited. Much larger sample sizes are needed to detect more modest effects (Figure 2B). Power may be further limited by disease heterogeneity, as histotype specific sample sizes are even smaller.

**Figure 2:**
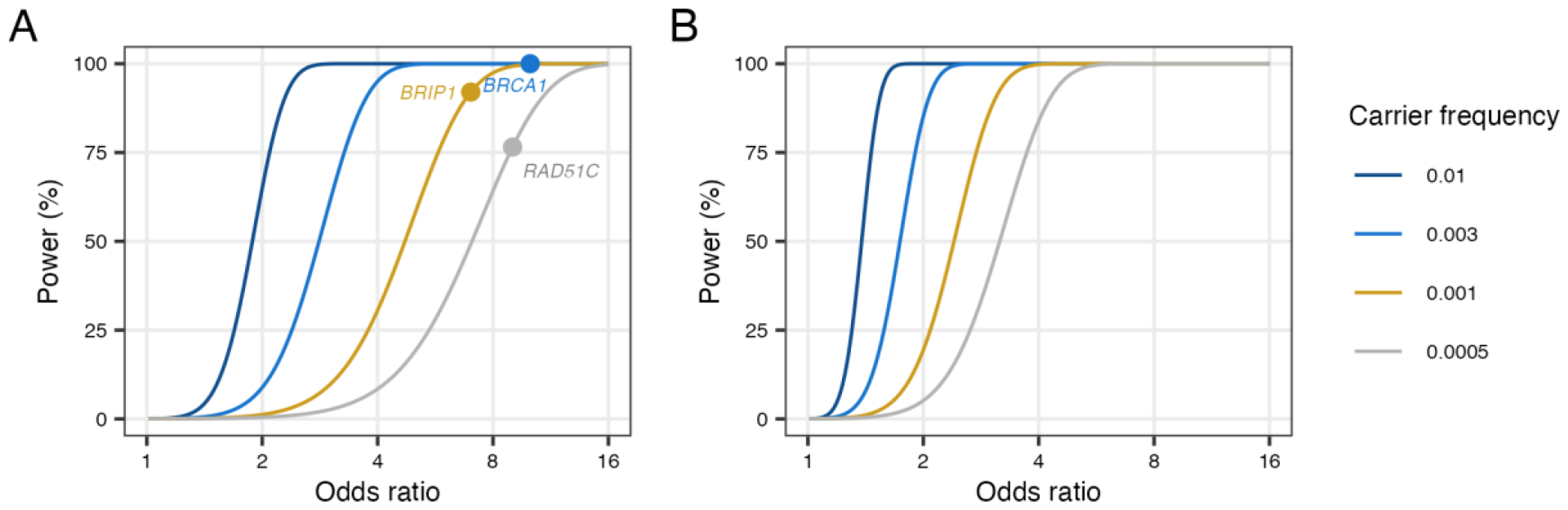
Power to detect risk alleles by carrier frequency and effect size (odds ratio) at a type 1 error probability of 5x10^-4^. A) 2,630 cases and 15,000 controls - the carrier frequency and effect size corresponding to BRCA1, BRIP1 and RAD51C are shown for reference. B) 20,000 cases and 20,000 controls

Nevertheless, we have confirmed the association of six genes known to be associated with high-grade serous ovarian cancer and have found evidence for the association with the same histotype for protein truncating variants in another three genes. However, the strength of the statistical evidence for these three genes is only moderate; while the Benjamini-Hochberg False Discovery Rate was less than 0.2, the Bayes False Discovery Probability was greater than 0.5. *NENF* encodes a neurotrophic factor that may play a role in neuron differentiation and development. In addition, circulating NENF levels have been shown to be decreased in subjects with polycystic ovary syndrome compared to controls ^26^. *MIGA1* encodes mitoguardin 1 which enables protein heterodimerization activity and protein homodimerization activity and is involved in mitochondrial fusion. The gene is expressed in the ovary and mitoguardin-1 and -2 promote maturation and the developmental potential of mouse oocytes by maintaining mitochondrial dynamics and functions ^27^. *STARD6* encodes the StAR related lipid transfer domain containing 6 protein which is involved in the intracellular transport of sterols and other lipids ^28^. It is notable that of the nine genes associated with high grade serous ovarian cancer three (*BRCA1, BRCA2, BRIP1*) were also associated with the non-high grade serous histotype (P<0.05) with another two (*NENF, RAD51D*) having a risk of the non-high grade serous histotype that was similar in magnitude but not statistically significant. Given the limited evidence for association of *BRCA1* and *BRCA2* with histoypes other than high-grade serous, this suggest some histotype misclassification in our data. There were too few pathogenic variant carriers in the non-high grade serous cases to estimate risk for the other four genes (*PALB2, RAD51C, MIGA1, STARD6*).

We have also confirmed the known association of the mis-match repair gene, *MSH6*, with the non-high grade serous histotype, with another four genes associated at a False Discovery Rate of less than 0.1. *HELB* encodes DNA helicase B which catalyzes the unwinding of DNA necessary for DNA replication, repair, recombination, and transcription ^29^ and rare damaging variants in the gene are associated with later age at natural menopause. Given the association of damaging variants with both age at natural menopause and non high grade serous ovarian cancer we used Mendelian randomisation to investigate the associations of genetically determined age at natural menopause with ovarian cancer by histotypes. Genome wide association studies have identified 290 common genetic variants associated with age at natural menopause ^30^. Summary statistics for the association of 234 of these variants with epithelial ovarian cancer by histotype were available to use as the instrumental variable. We applied Mendelian randomisation using five different methods implemented in the R package *TwoSampleMR* (MR Egger, weighted median, inverse variance weighted, simple mode and weighted mode). Five methods were used because each method is susceptible to different possible biases and consistent finding using different methods provides stronger evidence for any observed association A strong association was observed for endometrioid ovarian cancer (P < 0.05 for all five methods, Supplementary Table 3), with limited evidence for clear cell ovarian cancer (P < 0.05 for two methods) and little evidence for the other histoypes. Four of the six ovarian cases that were found to carry a loss-of-function variant in *HELB* were the endometriod histotype, with the other two being low-grade serous. Furthermore we analysed the data from whole-genome sequencing of tumour DNA from 59 high-grade serous, 35 clear cell and 29 endometrioid ovarian cancers ^31^ for point mutations in *HELB*. Only one pathogenic variant was identified in one of the endometrioid cases. The histotype specificity of the germline and somatic association of protein truncating variants in *HELB* together with the histotype specificity of the genetically predicted age at natural menopause association provides strong evidence that the association of protein truncating variants in *HELB* with non-high grade serous ovarian cancer is a true positive association.

*NBN* is also a good candidate ovarian cancer susceptibility gene. It encodes nibrin, a member of the MRE11/RAD50 double-strand break repair complex involved in DNA double-strand break repair and DNA damage-induced checkpoint activation. Protein truncating variants in this gene are associated with Nijmegen breakage syndrome, an autosomal recessive condition characterized by microcephaly, growth retardation, immunodeficiency, cancer predisposition, and premature ovarian failure in females ^32^. NBN has previously been studied using candidate-gene sequencing and no significant association was found for non-high grade serous ovarian cancer based on 444 non-high grade serous cases of which just 72 were the endometrioid histotype ^1^.

There is little evidence to link the *OR2T35* or *MYO1A* to the biology of ovarian cancer. *OR2T35* encodes olfactory receptor family 2 subfamily T member 35 and *MYO1A* encodes myosin 1A, an unconventional myosin that functions as actin-based molecular motors. This suggests the observed association with non high-grade serous ovarian cancer are likely to be false positives.

We have confirmed the histotype specific associations of rare protein truncating variants in the known epithelial ovarian cancer susceptibility genes and found two novel associations for protein truncating variants in *HELB* and *NBN* with risk of non-mucinous, non-high grade serous ovarian cancer. The relative risk estimates for these genes are likely to be inflated by the winners curse effect and may also be biased by the case ascertainement. Large case-control sequencing studies will be needed to obtain more precise, unbiased estimates of the associated risks as well as to obtain more specific risks for the three main histotypes that comprise non-mucinous, non-high grade serous ovarian cancer. Given our data it is unlikely that any additional susceptibility genes exist for either epithelial ovarian cancer of all histotypes or high-grade serous ovarian cancer with the risk-allele frequency and effect-size characteristics of the known susceptibility genes. It is possible there are genes with very rare risk alleles or modest effect sizes or genes specifically associated with the less common histotypes that we have not identified. Much larger studies will be needed to identify robustly such genes.

## Funding and acknowledgements

Cancer Research UK (PRCPJT-May21\100006); Cancer Australia Priority-driven Collaborative Cancer Research Scheme (APP1147276 to SJR); NIH (P30CA015083, P50CA136939, R01CA178535, R01CA248288 and R00CA256519). TD is supported by the German Research Foundation (Do761/15-1); SJR is supported by National Health and Medical Research Council (NHMRC) of Australia (2009840). Toon Van Gorp is a Senior Clinical Investigator of Research Foundation-Flanders (FWO)(18B2921N). The contents of the published material are solely the responsibility of the authors and do not reflect the views of the Caner Research UK, NHMRC, NIH, and other funders.

We thank: all the patients who have participated in the contributing studies and the many staff involved in participant recruitment; the NIHR Biomedical Research Centre at the University of Cambridge; the staff of the sequencing core at Ramaciotti Centre for Genomics (UNSW Sydney, Australia); Robert Geffers and the staff of the sequencing core at the Helmholtz Center HZI (Braunschweig, Germany).

## Ethics approval

The Institutional Review Board of Mayo Clinic, the East of England - Cambridge South Research Ethics Committee of UK National Research Ethics Service, the Institutional Review Board of Roswell Park Comprehensive Cancer Center, the Institutional Review Board of University of Southern California Health Sciences Campus, the Human Research Ethics Advisory Panels A and D of The University of New South Wales, Ethics Committee of Hannover Medical School, and Commissie voor Medische Ethiek/Klinisch Onderzoek of University of Leuven gave ethical approval for this work

## Data availability

We are unable to post the sequencing data due to ethical and/or legal data governance contraints on the sharing of Personal Data for some of the constituent studies.

## Author contributions

Conceptualization: SAG, PDPP, SJR

Methodology: EMD, SE, SAG, MJ, JB, P-CP, PDPP, JPT

Data assembly and analysis: EMD, MD, MP, P-CP, JPT

Resources: DB, JDB, IC, TD, ADF, EG, SAG, TVG, KM, KO, PDPP, SJR

Data curation: ED, MP, JPT

Writing - original draft: JB, EMD, SE, SAG, MJ, P-CP, PDPP

Writing – review and editing: All authors commented on the initial drafts of the manuscript and approved the final version.

Supervision: SAG, PDPP

Funding acquisition: SAG, PDPP, SJR

## Notes

### Competing Interest Statement

The authors have declared no competing interest.

### Author Declarations

East of England Cambridge South Research Ethics Committee of the UK National Research Ethics Service and the Institutional Review Board of University of Southern California Health Sciences Campus gave ethical approval for this work

